# The Diagnostic and Triage Accuracy of the GPT-3 Artificial Intelligence Model

**DOI:** 10.1101/2023.01.30.23285067

**Authors:** David M Levine, Rudraksh Tuwani, Benjamin Kompa, Amita Varma, Samuel G. Finlayson, Ateev Mehrotra, Andrew Beam

## Abstract

**Importance:** Artificial intelligence (AI) applications in health care have been effective in many areas of medicine, but they are often trained for a single task using labeled data, making deployment and generalizability challenging. Whether a general-purpose AI language model can perform diagnosis and triage is unknown.

**Objective:** Compare the general-purpose Generative Pre-trained Transformer 3 (GPT-3) AI model’s diagnostic and triage performance to attending physicians and lay adults who use the Internet.

**Design:** We compared the accuracy of GPT-3’s diagnostic and triage ability for 48 validated case vignettes of both common (e.g., viral illness) and severe (e.g., heart attack) conditions to lay people and practicing physicians. Finally, we examined how well calibrated GPT-3’s confidence was for diagnosis and triage.

**Setting and Participants:** The GPT-3 model, a nationally representative sample of lay people, and practicing physicians.

**Exposure:** Validated case vignettes (<60 words; <6^th^ grade reading level).

**Main Outcomes and Measures:** Correct diagnosis, correct triage.

**Results:** Among all cases, GPT-3 replied with the correct diagnosis in its top 3 for 88% (95% CI, 75% to 94%) of cases, compared to 54% (95% CI, 53% to 55%) for lay individuals (p<0.001) and 96% (95% CI, 94% to 97%) for physicians (p=0.0354). GPT-3 triaged (71% correct; 95% CI, 57% to 82%) similarly to lay individuals (74%; 95% CI, 73% to 75%; p=0.73); both were significantly worse than physicians (91%; 95% CI, 89% to 93%; p<0.001). As measured by the Brier score, GPT-3 confidence in its top prediction was reasonably well-calibrated for diagnosis (Brier score = 0.18) and triage (Brier score = 0.22).

**Conclusions and Relevance:** A general-purpose AI language model without any content-specific training could perform diagnosis at levels close to, but below physicians and better than lay individuals. The model was performed less well on triage, where its performance was closer to that of lay individuals.

## INTRODUCTION

Many artificial intelligence (AI) applications in health care have been shown to have high-accuracy and improve outcomes across a range of clinical applications including safety^1,2^, quality, and diagnosis.^3–6^ The vast majority of these AI systems are trained for a specific task (e.g., detecting solitary pulmonary nodules) using a single data modality (e.g., a database of chest CT scans). Moreover, such systems rely on human annotations^6,7^ which serve as the ground truth in the dataset. Labeling a large dataset is both labor-intensive^8^ and expensive. Health care workers must annotate a dataset for each individual task to be learned by the AI system. This single-task, single-model approach means that enormous effort is required to create an algorithm for a new task since new data must be acquired and labeled before the AI can learn to complete the task. Additionally, this process may only result in systems that will function optimally at the institution or with the dataset where it was trained^9–14^. This limits broader deployment of AI models in health care.

This barrier is being overcome outside of health care through general purpose, “self-supervised” AI models trained on generic tasks. These systems, often referred to as *foundation models*^15,16^, do not require labeled data in the traditional sense and have shown a remarkable ability to complete new tasks that they were not explicitly trained to perform^16–18^. For example, the Generative Pre-trained Transformer 3 (GPT-3) is a large self-supervised model that is trained only to predict the next word (i.e. “autocomplete”) using a large collection of unstructured text from the internet^17^. GPT-3 is one of the largest AI models ever created, containing over 175 billion model parameters, and was trained on approximately 570 gigabytes of data from the “common crawl” dataset, which contains nearly all unstructured text available on the Internet. Though GPT-3 was never trained for a specific task, GPT-3 has demonstrated the ability to answer questions, translate between languages, and converse interactively with people in a realistic manner^17^.

It is unknown if general AI models such as GPT-3 can provide reliable diagnostic information in health care.^9^ To date, empiric studies of GPT-3 in healthcare are limited to natural language processing tasks such as question answering and summarization.^21–23^ Recently, an updated version of GPT-3 known as “ChatGPT” has been released and has shown high-levels of accuracy on prep questions for medical licensing exams^19,20^. Though GPT-3 and ChatGPT share many components, ChatGPT’s simplified user interface allows for less control over the parameters that govern its responses and currently there is no publicly available API that would allow researchers to systematically study it. Given that, our focus is on GPT-3, though we anticipate that our results will likely generalize to ChatGPT.

If general AI models are successful in health care, this could lead to the rapid development of AI systems with little training, only validation. To advance our understanding of general AI models, we compared the diagnostic and triage accuracy of GPT-3 to lay individuals and primary care physicians. We focused on diagnosis and triage given this is a common task of professionals which requires understanding patient information provided in natural language.

## METHODS

### Design overview

We compared GPT-3’s triage and diagnostic performance to a nationally representative sample of lay Internet users and a sample of primary care physicians at Harvard Medical School. The Harvard Medical School IRB deemed this study not human subjects research.

### Setting and participants

We previously reported on the diagnostic and triage accuracy of adults who used the Internet^24^. Briefly, we enrolled participants using Toluna (Wilton, CT). We requested a nationally-representative sample of adults by gender, age, and census region. Participants were eligible if they were aged 18 years old or older and resided in the United States. Participants reviewed a simple case vignette, performed Internet searches to inform their thinking, and relayed their presumed diagnosis and triage regarding the case. Separately from the online participants, we previously enrolled a convenience sample of 21 primary care attending physicians at Harvard Medical School to validate the case vignettes. Physicians did not use the Internet.

### Patient vignettes and validation

Building on prior work evaluating symptom checkers,^13,14^ we previously^24^ created 48 case vignettes that included a chief complaint followed by additional pertinent details (**eTable 2**). Each vignette was less than 50 words and written at or below a 6^th^ grade reading level. Twelve vignettes were written for each of 4 triage categories: emergent, within 1-day, within 1-week, and self-care. We included both common (e.g., viral illness) and severe (e.g., heart attack) conditions but not highly obscure presentations. The correct diagnosis and triage category for each vignette were first determined by the authors (DML and AM, both general internists) and used as the gold standard. Engineers at OpenAI confirmed via correspondence that none of the vignettes used in our study appeared in GPT-3’s training data.

### Intervention and outcomes for humans

As previously described^24^, respondents (**eTable 1**), both lay people and physicians, were randomly assigned 1 of the 48 vignettes, asking them to “please read the following health problem, and imagine it were happening to your close family member.” After reviewing the vignette, they selected the triage they thought best: a) Let the health issue get better on its own. It most likely doesn’t require seeing a doctor; b) Try to see a doctor within a week. It likely won’t get better on its own, but it’s also not an emergency; c) Try to see a doctor within a day. The issue is urgent, but not an emergency; or d) Call 911 or go directly to the emergency room. The issue requires immediate attention. Next, participants answered the question, “What do you think are the three most likely medical diseases or diagnoses that could be causing this health problem?” This was a free-text response and respondents were asked to answer in order of likelihood.

Lay people were then asked to use the Internet in any way they believed to be useful to find the correct diagnosis and triage option for the health problem in the same vignette. After the Internet search, respondents reported the triage and diagnosis that they thought correct. For this analysis, we only used their second post-search responses.

### GPT-3 prompting procedure and parameters

We emulated the same process for GPT-3 as what the human participants performed; however, there were several key differences. For GPT-3 to complete a task, one must provide a “prompt” that will serve as the autocomplete template. For diagnosis prediction, we prompted GPT-3 using a fully complete vignette accompanied by the correct diagnosis and then asked GPT-3 to predict the diagnosis for a vignette it had not yet seen. Because GPT-3 is sensitive to how it is prompted, the model may return different diagnoses depending on which example case is used as a prompt. We prompted GPT-3 with each of the other 47 vignettes as examples and recorded its diagnosis and triage recommendation for each. Using this procedure, GPT-3 can provide a list of probable diagnoses. As with the humans, we marked a vignette correct if the ground truth diagnosis was among the top-3 most likely provided by GPT-3. See **eTable 3** for an analysis of the top-1 accuracy. To estimate GPT-3’s confidence on a given vignette, we divided the number of times each predicted diagnosis appeared in the prompting procedure by the total number of prompts (47 in this case) to produce a confidence score. The confidence score can be thought of as a pseudo-probability that ranges between 0 and 1, with a score of 1 indicating very high-confidence and 0 indicating very low-confidence.

We assessed accuracy of triage advice by examining whether the searcher’s selected triage was exactly correct (“exact”) according to the 4-level categorical scale above (emergent, 1-day, 1-week, and self-care) and whether the searcher’s selected triage matched a dichotomized triage variable (emergent/same-day vs 1-week/self-care; “dichotomized”). In our main analyses we present dichotomized triage, as we observed some inconsistency among physicians in triage with emergent vs. same-day^25^ (**eTable 4** for exact triage). We performed subgroup analysis by acuity. Using the same procedure as with diagnosis, we estimated confidence scores for GPT-3’s triage recommendations.

### Statistical analysis

We computed confidence intervals for performance measures using Wilson score intervals and compared performance between lay people, physicians, and GPT-3 using the Pearson *χ*^2^ test with continuity correction. We considered 2-sided p-values <0.05 to be significant. We performed all analyses using the R statistical computing language^26^, data and code to reproduce the results are available in the following github repository: https://github.com/beamlab-hsph/gpt3-clinical-vignettes.

To assess the quality of the confidence scores from GPT-3 we performed a calibration analysis^27,28^. Each confidence score was placed into one of five evenly placed bins (e.g. 0-20%, 20-40%, etc.) and the mean accuracy for the vignettes in each bin was computed. We created calibration curves for GPT-3’s diagnostic and triage confidence scores and additionally computed the Brier score^29^ as a global measure of calibration. A Brier score of 0 indicates perfect calibration while a score of 1 indicates complete miscalibration.

## RESULTS

### Diagnostic performance

Among all cases, GPT-3 was more likely to have the correct diagnosis in its top 3 for 88% (95% CI, 75% to 94%) of cases than lay individuals (54% (95% CI, 53% to 55%)) (p<0.001) though it was less likely to have the correct diagnosis compared to physicians 96% (95% CI, 94% to 97%) (p=0.03) (**Figure 2**). For self-care cases, GPT-3 and physicians had similar diagnostic performance: GPT-3 100% [95 % CI, 75% to 100%] vs physicians: 97.5% [95% CI, 94% to 99%], with lay individuals performing worse: 71% [95 % CI, 69% to 74%]. 1-week and 1-day cases had similar findings. Lay individuals again were less accurate than both GPT-3 and physicians: 44% correct on 1-day cases [95% CI, 39% to 44%] on 1-day cases and 59% on 1-week cases [95% CI, 56% to 62%]. For emergent cases, GPT-3 had an accuracy of 75% [95% CI, 55% to 95%], physicians had an accuracy of 94% [95% CI, 90% to 96%], and lay individuals had an accuracy of 43% [95% CI, 41% to 46%].

**Figure 1:**
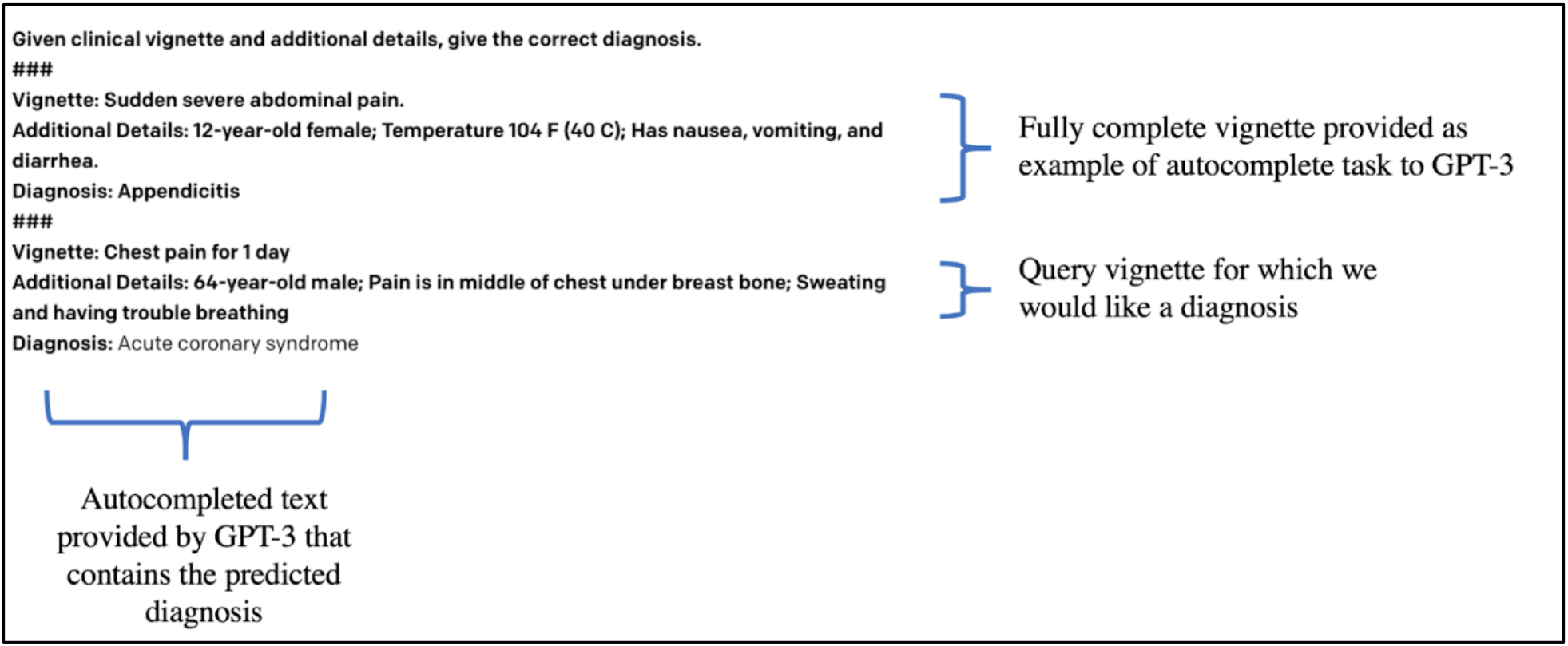
An illustrative example of GPT-3 prompting.

**Figure 2.**
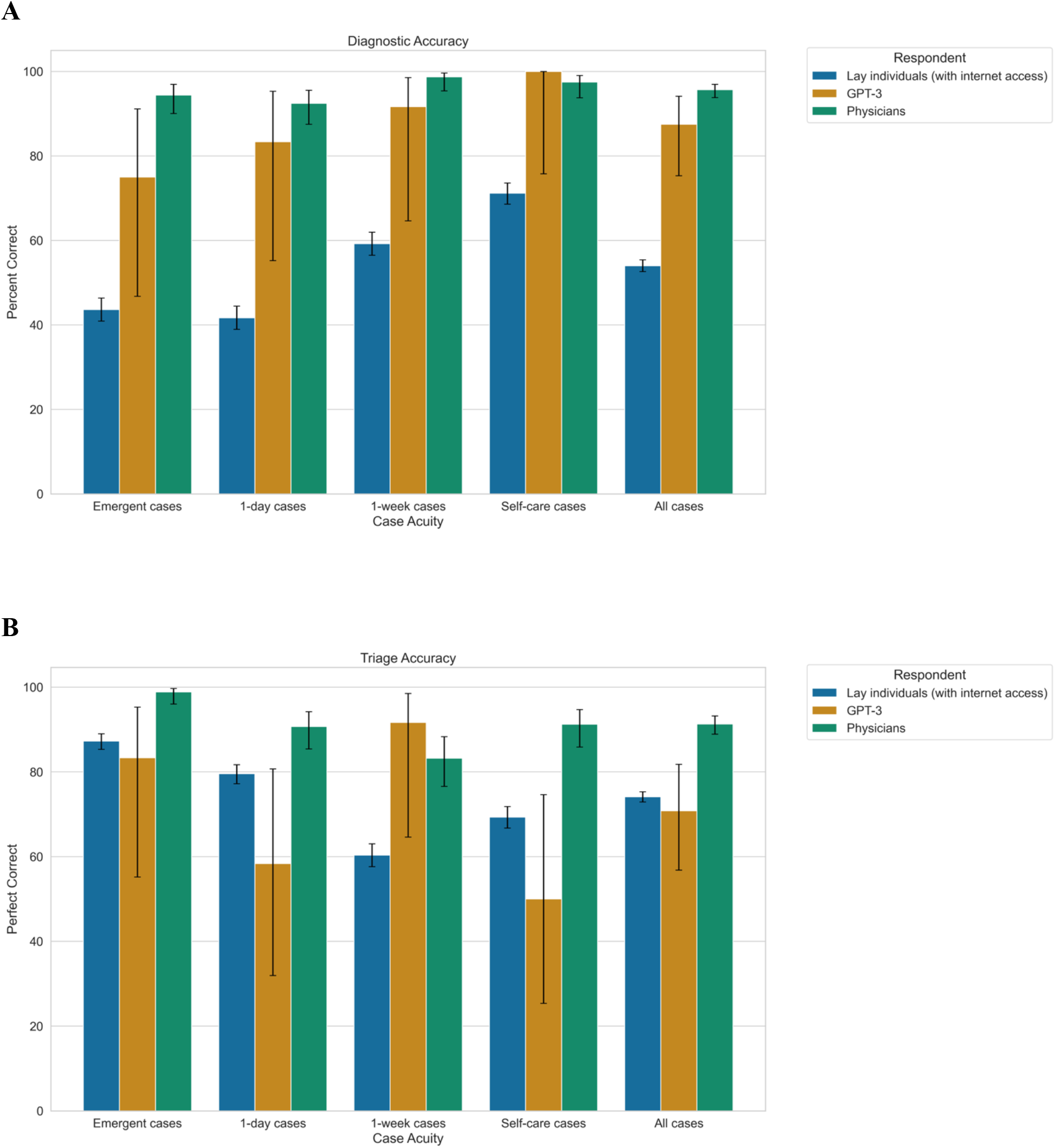
Diagnostic and triage accuracy of lay individuals with Internet access, GPT-3, and primary care attending physicians. **Panel A:** Correct diagnosis was listed among the top-3, stratified by case acuity from most acute (emergent) to least acute (self-care). **Panel B:** Correct dichotomized triage (same-day vs not same-day), stratified by case acuity from most acute (emergent) to least acute (self-care).

### Triage performance

Among all cases, GPT-3 had a triage accuracy of 70% [95% CI, 57% to 82%], which was similar to lay individuals (74%; 95% CI, 73% to 75%; p=0.73); both were significantly worse than physicians (91%; 95% CI, 89% to 93%; p<0.001). Performance varied by vignette severity. For example, GPT-3 performed slightly better than physicians for 1-week cases (92% [95 % CI, 65% to 99%] vs 83% [95% CI, 77%to 88%]) but performed worse on self-care cases (50% [95% CI, 25% to 75%].

### GPT-3 confidence and calibration

In general, GPT-3 had high-confidence in most of its predictions for both diagnosis and triage, and was more than 50% confident in its top prediction on the majority of cases. For diagnosis, the model had good calibration (Brier score = 0.18), though the calibration curve in Figure 3A reveals slight miscalibration for confidence scores < 50%, likely due to small sample sizes in these bins. A similar trend was observed for triage (Brier score = 0.22), with similar trends seen for triage predictions with low confidence.

**Figure 3.**
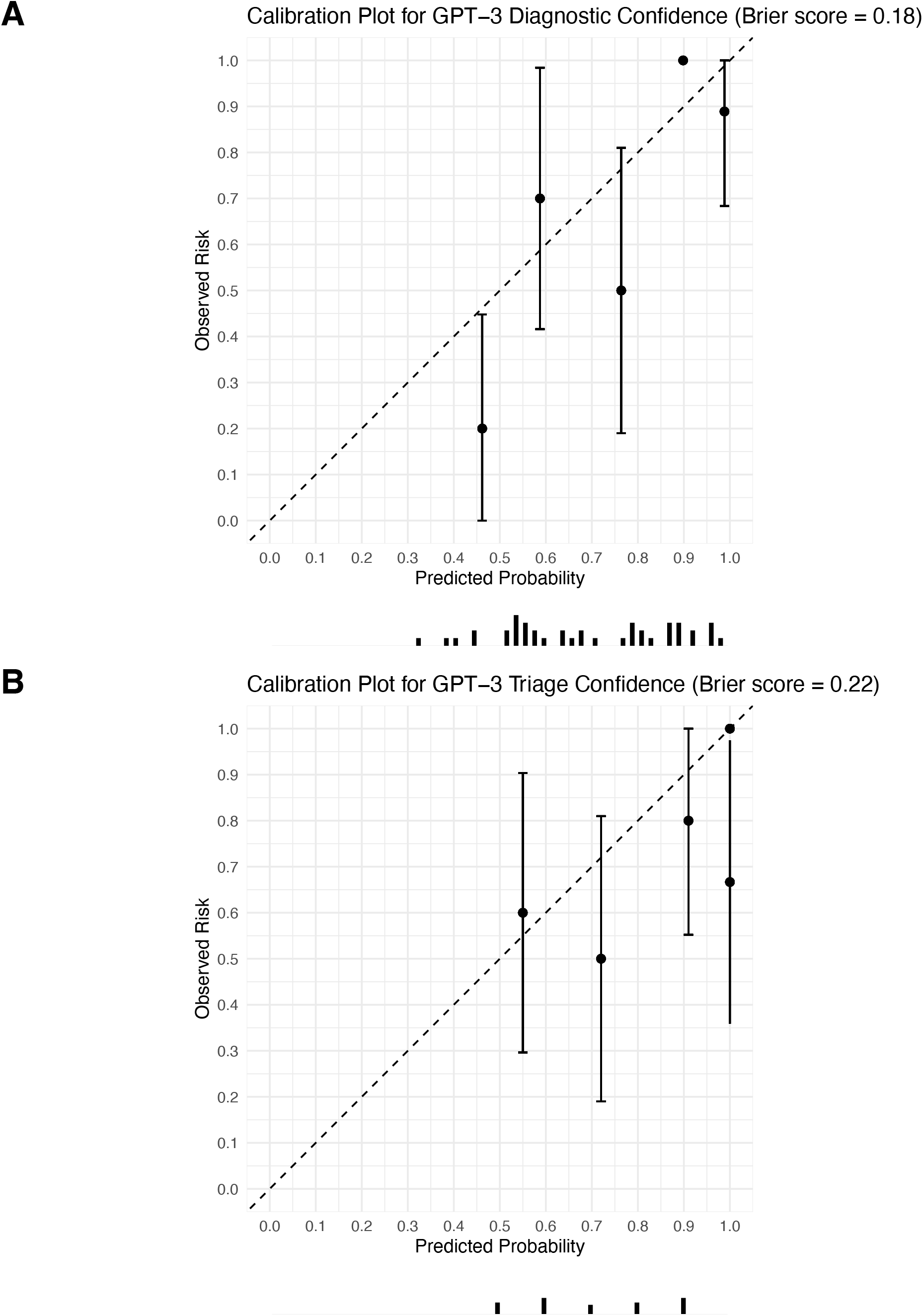
Calibration analysis of GPT-3 confidence scores for diagnosis and triage. Each panel shows a calibration curve displaying the relationship between GPT-3 confidence and accuracy. Each dot represents “binned” confidence scores that fall within the same range (e.g. 90-100%), while the y-axis shows the average accuracy for vignettes in that bin. In the case of good calibration, GPT-3 would get approximately 90% accuracy on vignettes that have a confidence score of approximately 90%. The Brier score is an overall summary of the calibration curve that ranges between 0 and 1, with a score of 0 implying perfect calibration. **Panel A**: Calibration curve and Brier score for GPT-3 diagnostic confidence. Overall, the model appears reasonably well-calibrated (Brier score=0.18). **Panel B**: Calibration curve and Brier score for triage prediction, again displayed evidence of reasonably good calibration (Brier score=0.22).

## DISCUSSION

In what we believe is the first evaluation of a generic AI tool’s performance in diagnosis and triage, we find that GPT-3’s performance was close to the performance of physicians for diagnostic accuracy, but inferior for triage. Its performance was much higher on both diagnosis and triage compared to lay people.

GPT-3’s diagnostic accuracy is notable given it was never trained explicitly to perform diagnosis or triage, nor was it trained using any kind of specialized medical data or patient records but instead was trained on a large corpus of text curated from the Internet^17^. It is possible that training on medical texts or triage guidelines could further improve performance beyond that of physicians. GPT-3 performed less well on triage. GPT-3’s learning from non-medical sources such as message boards may be part of the problem, and it is possible that adding triage manuals and other medical text to its library could improve its triage capability. It is also important to acknowledge that there is substantial variation among clinicians in the triage recommendation for these vignettes, as is the case for triage in general.

In the future, general AI tools such as GPT-3 could be used by patients and physicians to assist with diagnosis and triage. One advantage of such tools is GPT-3 is easy to use for untrained individuals and does not require pre-processing. For example, it may be possible for GPT-3 to “read” nurse or physician notes and generate diagnoses and triage recommendations or could support physicians and nurses in order entry, documentation, and other language-based tasks. Or perhaps patients could one day orally describe their concern and an automated transcript could be “read” by GPT-3 with resulting diagnosis and triage recommendations.

Our study advances prior research on general language models (which include models such as GPT-3) in health care and the use of GPT-3 specifically for health-related tasks.There is a large body of literature demonstrating language models such as GPT-3 result in significant improvements for pre-processing biomedical text^2,31–35^ and for prediction of important outcomes like disease onset^12,36^, mortality^37^, and readmission^38^. Recent work^39^ has shown that GPT-3 can reliably extract important clinical information such as medication mentions.

Despite GPT-3’s strong performance, there are numerous concerns with use of AI based on large language models. These kinds of AI models trained on large volumes of Internet text could amplify existing biases contained in the source data. Language models such as GPT-3 have been shown to demonstrate racial and gender bias^40,41^, and these biases could be difficult or impossible to correct as medical knowledge evolves^41^. Another example we produced that was outside of the scope for the present study was when asked about vaccines, GPT-3 responds, “Vaccines are not 100% effective. Vaccines can cause serious side effects. Vaccines can cause death. Vaccines are not tested for safety or effectiveness” (see supplement). When prompted with a query about the safety of smoking during pregnancy, one response GPT-3 offered was “Yes, it is safe to smoke during pregnancy. If you’re pregnant, you may be more likely to smoke as part of an effort to control your weight or because you are worried about quitting”. This is likely the result of GPT-3 being trained on text from the Internet, where misinformation can be rampant^42^. Guardrails must be in place before they can be safely used with patients and health care providers. An updated version known as “ChatGPT” was introduced in attempt to address some of these limitations. This model has incorporated human feedback and there are early anecdotes that this results in less toxic and more reliable behavior.

### Limitations

This study has limitations. First, our vignettes, while validated, are simulated cases. GPT-3, lay individuals, and physicians may perform differently when presented with real-world symptoms. Second, the manner in which GPT-3 is prompted can affect its output^43^. In this study we chose a straight-forward method of providing GPT-3 with an example vignette as a prompt, but it is possible that this type of prompt may not be available in all scenarios or to all potential users. Further work is needed to understand general prompting strategies for models such as GPT-3 that can be used in all applicable scenarios.

## CONCLUSIONS

The diagnostic accuracy of a generic AI model not specific to health care was higher than lay individuals and close to the performance of physicians. A generic model may afford a large opportunity to new AI applications that can be rapidly deployed without the burden of intensive and specialized training.

## Data Availability

All data produced are available online at https://github.com/beamlab-hsph/gpt3-clinical-vignettes

## Acknowledgments

- The authors would like to thank Girish Sastry, Raul Puri, Bianca Martin, and Miles Brundage from OpenAI for their assistance in working with GPT-3 and questions relating to GPT-3’s training data.

## Author contributions

- Tuwani had full access to all of the data in the study and takes responsibility for the integrity of the data and the accuracy of the data analysis.
- Study concept and design: Beam and Levine.
- Acquisition, analysis, or interpretation of data: All authors.
- Drafting of the manuscript: Beam and Levine.
- Critical revision of the manuscript for important intellectual content: All authors.
- Statistical analysis: Tuwani.
- Administrative, technical, or material support: ??.
- Study supervision: Beam.

