## Supplementary material for "The Diagnostic and Triage Accuracy of the GPT-3 Artificial Intelligence Model": Online supplement

David M Levine, MD, MPH, MA^1,2*^

Rudraksh Tuwani, BS^3,4*^

Benjamin Kompa, MPhil^4,5^

Amita Varma, BS^3,4^

Samuel G. Finlayson, PhD^6^

Ateev Mehrotra, MD, MPH^7^

Andrew Beam, PhD^4,5^

**SUPPLEMENTARY MATERIAL**

**eTable 1**. Participant characteristics

|  | Internet Searchers  (n = 5000)**^a^** |
| --- | --- |
| **AGE**, mean years (95% CI) | 45.0 (44.5,45.4) |
|  | **n (%)** |
| **FEMALE** | 2549 (51.0) |
| **RACE/ETHNICITY** |  |
| Non-Hispanic White | 3819 (76.4) |
| Non-Hispanic Black | 404 (8.1) |
| Hispanic | 318 (6.4) |
| Non-Hispanic Asian | 309 (6.2) |
| Non-Hispanic Other or Multiple | 150 (3.0) |
| **CENSUS REGION** |  |
| Northeast | 950 (19.0) |
| Midwest | 1150 (23.0) |
| South | 1750 (35.0) |
| West | 1150 (23.0) |
| **PARTNERED** | 2848 (57.0) |
| **EDUCATION** |  |
| <High school | 103 (2.1) |
| High school/GED | 1047 (20.9) |
| Some college | 1709 (34.2) |
| Bachelor's degree | 1435 (28.7) |
| >Bachelor's | 706 (14.1) |
| **HEALTH INSURANCE COVERAGE** |  |
| Uninsured | 443 (8.9) |
| Medicare | 1147 (22.9) |
| Medicaid | 535 (10.7) |
| Both Medicare and Medicaid | 187 (3.7) |
| Private/employer-based | 2484 (49.7) |
| Not sure | 204 (4.1) |
| **PERCEIVED HEALTH STATUS** |  |
| Excellent | 690 (13.8) |
| Very good | 1780 (35.6) |
| Good | 1794 (35.9) |
| Fair | 605 (12.1) |
| Poor | 131 (2.6) |
| **EMPLOYED** | 2940 (58.8) |
| **FAMILY INCOME** |  |
| <$30,000 | 1260 (25.2) |
| $30,000-$49,999 | 991 (19.8) |
| $50,000-$79,999 | 1182 (23.6) |
| $80,000-$99,999 | 534 (10.7) |
| $100,000-$149,999 | 660 (13.2) |
| $150,000-$199,999 | 225 (4.5) |
| $200,000 or more | 148 (3.0) |
| **CHRONIC DISEASES** |  |
| 0 | 2612 (52.2) |
| 1 | 1066 (21.3) |
| 2 | 617 (12.3) |
| >2 | 520 (10.4) |
| Not sure | 185 (3.7) |
| **HAS PRIMARY CARE** | 3963 (79.3) |
|  | **mean (95% CI)** |
| **PHYSICIAN VISITS** in last 6 months | 2.1 (2.0,2.1) |
| **EMERGENCY ROOM VISITS** in last 6 months | 0.3 (0.3,0.3) |
| **HOSPITAL ADMISSIONS** in last 6 months | 0.2 (0.2,0.2) |
| **GLOBAL HEALTH CARE RATING** in last 6 months (0-10) | 7.4 (7.3,7.5) |

**^a^**	Nationally-representative sample by age, gender, and census region. Percents may not sum to 100 due to rounding.

**Note**: As reported in Levine and Mehrotra

**Abbreviations**: CI, confidence interval; GED, general educational development

**eTable 2**. Clinical case vignettes

| **Case #** | **Correct Diagnosis** | **Correct Triage** | **Current Problem** | **Additional Details** |
| --- | --- | --- | --- | --- |
| 1 | Liver failure | Emergent | Confused and sleepy for several hours | 48-year-old female  Can’t answer where she is or what her name is (but she normally can)  Takes Tylenol all the time for migraine, but recently took more |
| 2 | Appendicitis | Emergent | Sudden severe abdominal pain | 12-year-old female  Temperature 104 F (40 C)  Has nausea, vomiting, and diarrhea |
| 3 | Heart attack | Emergent | Chest pain for 1 day | 64-year-old male  Pain is in middle of chest under breast bone  Sweating and having trouble breathing |
| 4 | Kidney stones | Emergent | Left-sided pain between armpit and hip for 1 hour | 45-year-old male  Severe pain  Pain starts on left side and travels to groin  Has nausea and vomiting  Squirming in pain |
| 5 | Meningitis | Emergent | Headache for 3 days | 18-year-old male  Has fever 102  Neck stiff  Light bothers him |
| 6 | Pulmonary embolism | Emergent | Trouble breathing for 30 minutes | 65-year-old male  Has chest pain that worsens when he takes a breath  Had surgery last week  Has swelling and pain in his left calf |
| 7 | Rocky Mountain Spotted Fever | Emergent | Fever and rash for 2 days | 8-year-old male  Rash is worse on the ankles and wrists  Has joint pain and headache  Was camping recently |
| 8 | Stroke | Emergent | Right-sided weakness and difficulty speaking for 10 minutes | 70-year-old male  Can’t use his right arm  Has high blood pressure and abnormal heart rhythm  Has nausea and vomiting |
| 9 | Tetanus | Emergent | Painful muscle spasms for 1 day | 65-year-old male  Having a hard time opening his mouth  Cut himself while gardening  He is restless and irritable |
| 10 | COPD exacerbation | Emergent | Trouble breathing for 3 days | 67-year-old female  Has lung disease and smoked cigarettes  Increased cough with green phlegm  Can’t speak in complete sentences |
| 11 | Asthma exacerbation | Emergent | Trouble breathing for 3 days | 27-year-old female  Recent cold  Wheezing and coughing when sitting  Trouble breathing when walking around house  Has asthma  Inhalers don’t help her anymore |
| 12 | Heart failure exacerbation | Emergent | Trouble breathing for 3 days | 78-year-old female  Ate very salty foods recently  Gaining weight and legs are swollen  Had a heart attack several years ago  Trouble breathing when sitting on the couch |
| 13 | Cellulitis | 1 day | Front of left leg is red and hurts for one day | 45-year-old man  Redness started quickly over a day  He thinks he has a high temperature, but he didn’t check  Leg is swollen  Leg hurts to touch it, but he didn’t cut it or bump it that he remembers |
| 14 | Strep throat | 1 day | Fever and sore throat for 2 hours | 7-year-old female  White stuff in back of throat  Painful front of the neck  No cough, no congestion  Temperature 102 |
| 15 | Asthma exacerbation | 1 day | Trouble breathing for 3 days | 27-year-old female  Recent cold  Wheezing and coughing, especially at night  Has asthma  Inhalers only help for a couple of hours |
| 16 | COPD exacerbation | 1 day | Trouble breathing for 3 days | 67-year-old female  Has lung disease and smoked cigarettes  Increased cough with green phlegm  Using inhaler every 6 hours |
| 17 | Deep vein thrombosis | 1 day | Right leg pain for 2 days | 65-year-old female  Right leg is swollen, red, and painful  Was in hospital last week for pneumonia  Has mild heart disease |
| 18 | Hemolytic uremic syndrome | 1 day | Belly pain and diarrhea for 7 days | 4-year-old male  Nothing unusual in diet though did have a hamburger at a cookout 3 days before pain started  Has fever  Diarrhea may have blood in it |
| 19 | Malaria | 1 day | Fever and chills for 5 days | 18-year-old male  Has fever of 103.1F  Recently returned from Africa |
| 20 | Pneumonia | 1 day | Cough and fever for 3 days | 65-year-old male  Cough brings up green phlegm  Breathing fast and shallow |
| 21 | Mononucleosis | 1 day | Fever, sore throat, enlarged lymph nodes for one week | 16-year-old female  Trouble swallowing  Extremely tired |
| 22 | Salmonella | 1 day | Vomiting, diarrhea, and belly pain for 18 hours | 14-year-old male  Had diarrhea 10 times in 18 hours  No blood in diarrhea  Has temperature of 103.1F  Attended a picnic recently and ate undercooked chicken |
| 23 | Shingles | 1 day | Pain on right side of chest for 5 days | 77-year-old male  Pain was first and was then followed by rash in same area as pain  Feels unwell in general  Rash becomes clear pockets of fluid on top of skin after 3 days |
| 24 | Urinary tract infection | 1 day | Painful urination for 2 days | 26-year-old female  Has urgent need to pee  Recent sexual activity  No fever, but some back pain |
| 25 | Acne | 1 week | Red bumps on forehead for 4 weeks | 17-year-old male  Bumps come and go and then come in new places on face |
| 26 | Acute sinusitis | 1 week | Feels congested and face pain for 15 days | 35-year-old female  Has green nose discharge  Has a fever again after thinking she was getting better |
| 27 | Peptic ulcer disease | 1 week | Upper abdominal pain for 2 months | 40-year-old male  Feels like a dull, gnawing ache  Wakes him up at night  Better when drinking milk and eating |
| 28 | Vertigo | 1 week | Sudden episodes of dizziness for 1 month | 65-year-old female  Triggered by tilting her head back to look up  Each attack lasts about 30 seconds  No hearing problems or weakness |
| 29 | Diabetes | 1 week | Thirsty and peeing a lot for 4 weeks | 52-year-old male  Feels tired all the time  Has blurry vision on and off |
| 30 | Hemorrhoids | 1 week | Blood on toilet paper for 1 week | 68-year-old female  Stool is hard and must strain when using the bathroom  No weight loss, belly pain, or fever  Stool is not red or black |
| 31 | Osteoarthritis | 1 week | Left knee pain for 4 months | 62-year-old male  Worse after walking, better when resting  Sometimes swells up  Never hit knee against anything or fell |
| 32 | Depression | 1 week | Feeling down for weeks | 32-year-old male  Wakes up early and cannot go back to sleep  Having difficulty concentrating at work  Cannot seem to stop worrying about his family |
| 33 | Obstructive sleep apnea | 1 week | Tired for 2 months | 42-year-old female  Overweight  Falls asleep during the day  Partner complains about snoring at night |
| 34 | Heart failure exacerbation | 1 week | Trouble breathing for 7 days | 78-year-old female  Slowly gaining weight  Had a heart attack several years ago  Comfortable when sitting or walking slowly, but has trouble when walking up stairs |
| 35 | Hypothyroidism | 1 week | Tired for 2 months | 39-year-old female  Has dry skin, weight gain, and constipation  Feels very cold when others are hot |
| 36 | Gastroesophageal reflux disease | 1 week | Upper abdominal pain for 2 months | 40-year-old male  Worse with spicy and fried foods  Worse if he eats late at night before sleeping  Voice is becoming hoarse |
| 37 | Common cold | Self-care | Runny nose for 12 days | 34-year-old female  No fever  Had also had a sore throat and cough  No other medical problems |
| 38 | Viral conjunctivitis | Self-care | Red, irritated eye for 3 days | 14-year-old male  Spread from right to left  Has watery eye discharge, but no pain  Has a stuffy nose |
| 39 | Viral pharyngitis | Self-care | Sore throat for 2 days | 26-year-old male  Has a headache  Has a cough  No fever |
| 40 | Allergic rhinitis | Self-care | Congestion for many years | 22-year-old male  Worse during spring season  Has sneezing and nose itching  Has eye itching and tearing |
| 41 | Musculoskeletal low back pain | Self-care | Back pain for 3 weeks | 35-year-old male  Pain started after shoveling snow  No leg pain or weakness  No fever or weight loss |
| 42 | Bee sting without anaphylaxis | Self-care | Swollen and tender forehead at site of bee sting for 1 hour | 9-year-old male  Stopped crying after 15 minutes  Tongue looks normal  No wheezing or breathing problems |
| 43 | Canker sore | Self-care | Mouth sores that come back over several years | 17-year-old male  Has 5 of them in his mouth  No other sores anywhere else  Takes no drugs or medicines |
| 44 | Candidal yeast infection | Self-care | White stuff coming out of vagina for 2 days | 40-year-old female  Vagina also itchy  Doesn’t hurt to pee  No abdominal pain  No fever |
| 45 | Eczema | Self-care | Dry itchy skin in front of elbows and behind knees for years | 12-year-old female  Brother has asthma  Recently told she has egg and milk allergies |
| 46 | Stye | Self-care | Painful swollen right eyelid for 1 day | 30-year-old male  Pain is at edge of eyelid  Hurts to touch it  No change in vision |
| 47 | Influenza | Self-care | Fever, headache, and cough for 4 days | 30-year-old female  Came on very suddenly  Feels weak all over  Temperature 102.5 at first, but not now |
| 48 | Poison Ivy | Self-care | Itchy rash on left hand for 1 day | 12-year-old male  Came on hours after playing in woods  Rash red and warm  No fever |

**eTable 3: GPT-3 Top-1 Accuracy**

|  | **N** | **# Correct** | **Mean** | **N** | **95% CI Lower** | **95% CI Upper** |
| --- | --- | --- | --- | --- | --- | --- |
| **1-day** | 12.0 | 7.0 | 58.333333 | 12.0 | 31.951131 | 80.673969 |
| **1-week** | 12.0 | 8.0 | 66.666667 | 12.0 | 39.062209 | 86.187991 |
| **Emergent** | 12.0 | 8.0 | 66.666667 | 12.0 | 39.062209 | 86.187991 |
| **Self-care** | 12.0 | 8.0 | 66.666667 | 12.0 | 39.062209 | 86.187991 |
| **All** | 48.0 | 31.0 | 64.583333 | 48.0 | 50.439057 | 76.566356 |

**eTable 4: GPT-3 Exact Triage Accuracy**

|  | **N** | **# Correct** | **Mean** | **95% CI Lower** | **95% CI Upper** |
| --- | --- | --- | --- | --- | --- |
| **1-day** | 12.0 | 2.0 | 16.666667 | 4.696514 | 44.803086 |
| **1-week** | 12.0 | 11.0 | 91.666667 | 64.612009 | 98.513491 |
| **Emergent** | 12.0 | 6.0 | 50.000000 | 25.378160 | 74.621840 |
| **Self-care** | 12.0 | 2.0 | 16.666667 | 4.696514 | 44.803086 |
| **All** | 48.0 | 21.0 | 43.750000 | 30.701269 | 57.724983 |

**eAppendix: Details of GPT-3 prompting procedure**

Our template prompts for diagnosis prediction is available on <https://beta.openai.com/playground/p/CmtySXvhwmij1uJT2bM02Wka?model=davinci-instruct-beta> and those for triage prediction is available on <https://beta.openai.com/playground/p/KdDrTEzsDmOXJt0m4dbUQV3x?model=davinci-instruct-beta>. Both the templates require access to the GPT-3 API.

**GPT-3 Parameters for diagnosis and triage prediction**

| Name | Value |
| --- | --- |
| Engine | davinci-instruct-beta |
| Temperature | 0 |
| Top P | 1 |
| Frequency Penalty | 0 |
| Presence Penalty | 0 |
| Best Of | 1 |
| Stop Sequences | ‘###’, ‘\n’ |

**Example diagnosis prompt:** (<https://beta.openai.com/playground/p/CmtySXvhwmij1uJT2bM02Wka?model=davinci-instruct-beta>):

Given clinical vignette and additional details, give the correct diagnosis.

\###

Vignette: Sudden severe abdominal pain.

Additional Details: 12-year-old female; Temperature 104 F (40 C); Has nausea, vomiting, and diarrhea.

Diagnosis: Appendicitis

\###

Vignette: Confused and sleepy for several hours.

Additional Details: 48-year-old female; Can't answer where she is or what her name is (but she normally can); Takes Tylenol all the time for migraine, but recently took more.

Diagnosis:

**Example triage prompt:** (<https://beta.openai.com/playground/p/KdDrTEzsDmOXJt0m4dbUQV3x?model=davinci-instruct-beta>):

Given a clinical vignette and additional details, classify into one of the 4 triage categories: 1-day, 1-week, Emergent, and Self-care.

\###

Vignette: Painful urination for 2 days.

Additional Details: 26-year-old female; Has urgent need to pee; Recent sexual activity; No fever, but some back pain.

Triage: 1-day

\###

Vignette: Feeling down for weeks.

Additional Details: 32-year-old male; Wakes up early and cannot go back to sleep; Having difficulty concentrating at work; Cannot seem to stop worrying about his family.

Triage:
